# Individual-level Structural Neuroimaging Biomarkers across the Psychosis Spectrum

**DOI:** 10.64898/2026.10.18.26365847

**Authors:** Alexa M. Rivas, Efe Sari, Cemal Demirlek, Akila Weerasekera, Abigail Stein, Jonah Loshin, Virginie-Anne Chouinard, Mark Halko, Roscoe O Brady, Kathryn Eve Lewandowski, Michael Murphy, Ann K. Shinn, Cagri Yuksel, Zachary B. Millman, Fei Du, Marek Kubicki, Dost Öngür, Halide Bilge Türközer

## Abstract

**Background and Hypothesis:** Although psychotic disorders are characterized by widespread cortical thinning, translating these findings into individualized biomarkers remains challenging. Regional Vulnerability Index (RVI) and Neuroscore are candidate individual-level biomarkers that quantify the similarity between an individual’s structural brain pattern and generic psychosis-associated alterations. This study investigated cortical thickness (CT)-based RVI and Neuroscore, along with mean CT deviation from healthy individuals, across the psychosis spectrum.

**Study Design:** Using structural 3T MRI data from 289 participants (clinical high risk for psychosis [CHR], n=39; first episode psychosis [FEP], n=44, chronic psychosis [CP], n=86; healthy controls [HC], n=120), RVI, Neuroscore, and mean CT deviation (z) scores were computed from region-wise CT measures quantified using FreeSurfer v7.2. Group differences and associations with symptoms were evaluated.

**Study Results:** RVI and Neuroscore were significantly higher, and mean CT z-scores lower, in FEP and CP compared with CHR and HC, with moderate-to-large effect sizes (|Cohen’s d| = 0.55–1.15). No differences were observed between FEP and CP or CHR and HC. Indices were not associated with psychosis-spectrum symptom severity in the CHR, FEP, or CP groups. RVI (*r* = −0.561, *p* < .001) and Neuroscore (*r* = −0.997, *p* < .001) were strongly negatively correlated with the mean CT z-score.

**Conclusions:** CT-based RVI and Neuroscore appear to index established disease status but not risk status, while showing substantial overlap with mean CT deviation. These findings highlight the need to further refine these biomarkers through data-driven and multimodal approaches to improve their utility for risk and prognostic prediction.

## INTRODUCTION

Schizophrenia Spectrum Disorders (SSD) are a leading cause of psychiatric disability, often contributing to significant functional impairment, lifelong treatment, and poor recovery outcomes.^1^ Despite this, well-established predictors of psychosis risk and prognosis to enable early intervention are limited, with current approaches demonstrating only modest predictive accuracy and substantial heterogeneity across individuals.^2–4^ Accurately identifying individuals at risk of developing psychosis and those experiencing poor clinical outcomes remains a critical goal for achieving effective early intervention and improving treatment outcomes.

Despite the large body of research showing various neurobiological alterations in SSD^5^, applying these group-level findings to individual-level risk assessment and prognostic prediction has been challenging, limiting clinical utility. These limitations have fueled growing interest in exploring biomarkers that can capture variation within individuals rather than between diagnostic groups. Earlier efforts focused on normative models, which quantify deviations from brain measures observed in the general population.^6–8^ Recent work has focused on translating meta-analytic findings in SSD into individual-level biomarkers. The Regional Vulnerability Index (RVI) and Neuroscore are examples of this approach, which quantify the degree to which an individual’s structural brain alterations resemble those observed in SSD.^9,10^ The RVI quantifies the similarity between an individual’s structural brain profile and the characteristic pattern of schizophrenia-related abnormalities derived using effect sizes from the large-scale ENIGMA (Enhancing NeuroImaging Genetics through Meta-Analysis) dataset.^9^ Similarly, the Neuroscore applies ENIGMA-derived effect size weights to regional brain measures and aggregates the weighted values into a single summary score.^10^ These indices have shown associations with clinical characteristics in prior SSD studies. RVI has shown to be associated with negative symptoms in psychosis ^9^ and familial psychosis loading in children with ancestral family history of SSD^11^, while Neuroscore has been associated with both positive and negative symptom severity in psychosis.^10,12^

Although these approaches have been instrumental in bringing an individual-level perspective to psychosis research, critical gaps remain. First, there is limited exploration of these biomarkers across the full psychosis spectrum from clinical high risk for psychosis (CHR) to first episode psychosis (FEP) and chronic psychosis (CP). This is essential to determine how these measures evolve across illness stages and relate to clinical features. Second, direct comparisons of different types of individual-level biomarkers are limited, leaving uncertainty regarding whether they differ in their ability to capture clinically relevant variation and their potential clinical utility.

The present study addresses these gaps by evaluating cortical thickness-based RVI and Neuroscore across the psychosis spectrum, including CHR, FEP, and CP. Additionally, we quantified a mean cortical thickness (CT) standardized deviation score (mean CT z-score) to assess overall cortical thickness deviation from healthy controls alongside RVI and Neuroscore. We compared these three biomarkers across CHR, FEP, CP, and healthy control (HC) groups and examined their associations with psychosis-spectrum symptom severity in the CHR, FEP, and CP groups, as well as their relationships with one another. By characterizing these biomarkers across illness stages and evaluating their clinical correlates, this study advances the development and refinement of individualized neuroimaging biomarkers for risk prediction and prognostic characterization in psychotic disorders.

## METHODS

### Participants

Clinical and imaging data from studies conducted within the McLean Psychotic Disorders’ Division were used for this study.^13–15^ Individuals were recruited from inpatient and outpatient hospital services and via advertisement for these studies. All included studies were approved by the Institutional Review Board, and written informed consent was obtained from all participants. The final sample included 289 individuals (HC, n = 120; CHR, n = 39; FEP, n = 44; CP, n = 86). Individuals with psychotic disorders (schizophrenia, schizoaffective disorder, or schizophreniform disorder) and a disease duration shorter than 3 years were classified as FEP, whereas those with disease duration greater than 3 years were classified as CP.^16^ Disease duration was calculated as the difference between current age and age of onset. See **Table 1** for demographic and clinical characteristics.

**Table 1.**
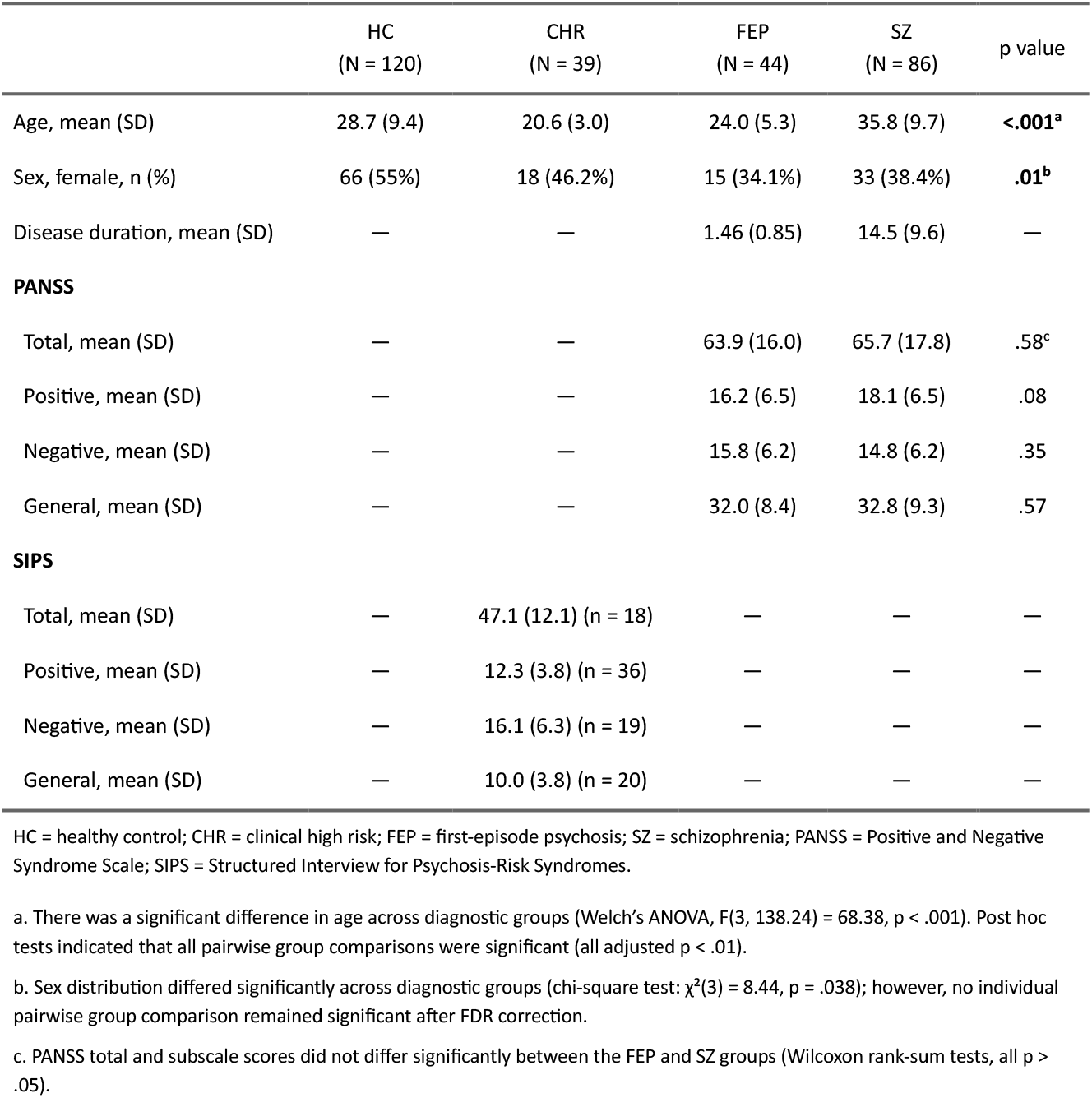
Demographic and clinical characteristics of cohort.

### Clinical Assessments

Psychiatric diagnoses were assessed using the Structured Clinical Interview for DSM-V (SCID-5). Structured Interview for Psychosis Risk Syndromes (SIPS) was used to determine CHR status.^17^ SIPS positive, negative, general, and total symptom scores were used to assess psychosis-spectrum symptoms in the CHR group. For individuals in the FEP and CP groups, psychotic symptom severity was assessed using the Positive and Negative Syndrome Scale (PANSS), including positive, negative, and general symptom subscales.

### Imaging Processing

The study utilized structural neuroimaging scans acquired using two 3T MRI scanners (Siemens Trio and Siemens MAGNETOM Prisma, **Table S1 and S2**) with a standardized 3D MPRAGE sequence. Scans were processed using FreeSurfer’s automated pipeline (v.7.2).^18^ Cortical thickness values for 34 bilateral Desikan-Killiany (DK) atlas regions were quantified.^19^ All outputs were visually inspected for motion artifacts and to ensure proper gray/white matter segmentation. Outliers were examined, and CT measures were winsorized at three standard deviations (SD) from the mean. In total, 35 participants had at least one outlying CT value, comprising 67 individual left- or right-hemisphere regional CT measures.

### Indices (RVI, Neuroscore, mean CT z-score)

RVI and Neuroscore were calculated following methods described in Kochunov et al.^9^ and Rodrigue et al.^10^ respectively. For each cortical region, CT values from the left and right hemispheres were averaged, yielding 34 bilateral mean CT measures. Age, sex, and scanner effects were regressed from the regional CT measures, and the resulting unstandardized residuals were saved. This approach deviated from the original RVI methodology by not adjusting for intracranial volume (ICV) or race, given FreeSurfer’s recommendation against ICV adjustment for CT measures and growing evidence cautioning against the use of race as a biological covariate in neuroimaging analyses.^20^ Residualized CT measures were then standardized using the mean (M) and standard deviation (SD) derived from healthy controls.

The CT-based RVI was calculated as the Pearson correlation coefficient between each individual’s 34 region-wise CT z-scores and the corresponding effect sizes for CT differences between schizophrenia and control groups derived from the ENIGMA meta-analysis.^5^ Higher RVI values indicate greater similarity between an individual’s spatial pattern of cortical thickness variation and the characteristic pattern of schizophrenia-associated cortical thickness abnormalities identified by ENIGMA.

The CT-based Neuroscore was calculated using 68 region-wise CT z-scores, with left- and right-hemisphere measures retained separately. For each individual, each regional CT z-score was multiplied by the corresponding effect size from the same ENIGMA meta-analysis ^5^, and the resulting weighted regional values were summed to generate a single Neuroscore. Higher Neuroscore values indicate greater overall similarity to the schizophrenia-associated cortical thickness alterations reported by ENIGMA.

A mean CT standardized deviation score (mean CT z-score) was also calculated for each individual by averaging the 34 region-wise CT z-scores. Lower values indicate greater overall negative deviation, whereas higher values indicate greater overall positive deviation, relative to CT values observed in HC.

Although scanner effects were regressed out from regional CT measures prior to index calculation, we additionally assessed the final RVI, Neuroscore, and mean CT z-score values across scanners to ensure that no residual scanner differences remained; no significant differences were observed within the FEP, HC, or CP groups (**Table S3**). CHR data were acquired on a single scanner.

### Statistical Analysis

All statistical analyses were conducted in RStudio version 2024.04.2+764. Outliers were assessed for RVI, Neuroscore, and mean CT z-scores, using a threshold of ±3 standard deviations (SD) from the mean; no observations exceeded this threshold and all were retained for analysis. The three indices and clinical variables were normally distributed except for PANSS subscale and total scores. Group differences in each index across CHR, FEP, CP, and HC groups were assessed using one-way analyses of variance (ANOVAs). Significant effects were followed by Tukey’s honestly significant difference (HSD) post hoc tests, and pairwise effect sizes were quantified using Cohen’s d. Correlations among RVI, Neuroscore, and mean CT z-scores were assessed using Pearson correlations. The indices’ associations with SIPS scores were examined using Pearson correlations in the CHR group, whereas associations with PANSS scores and disease duration were assessed using Spearman rank correlations in the FEP and CP groups due to their non-normal distributions. The false discovery rate (FDR) method was applied to correct for multiple comparisons.^21^ Corrections were applied separately across families of tests comprising group comparisons, correlations among the indices, correlations with SIPS scores in CHR, correlations with PANSS scores in FEP and CP groups, and correlations with duration of illness. FDR-corrected p (q) values were reported, with q < 0.05 considered statistically significant.

## RESULTS

### Group differences

Omnibus statistics for all three indices showed significant differences across the four groups (RVI, F(3, 285) = 14.36, *q* < .001; Neuroscore, F(3, 285) = 17.8, *q* < .001; mean CT z-score, F(3, 285) = 16.42, *q* < .001) (**Figure 1**).

**Figure 1.**
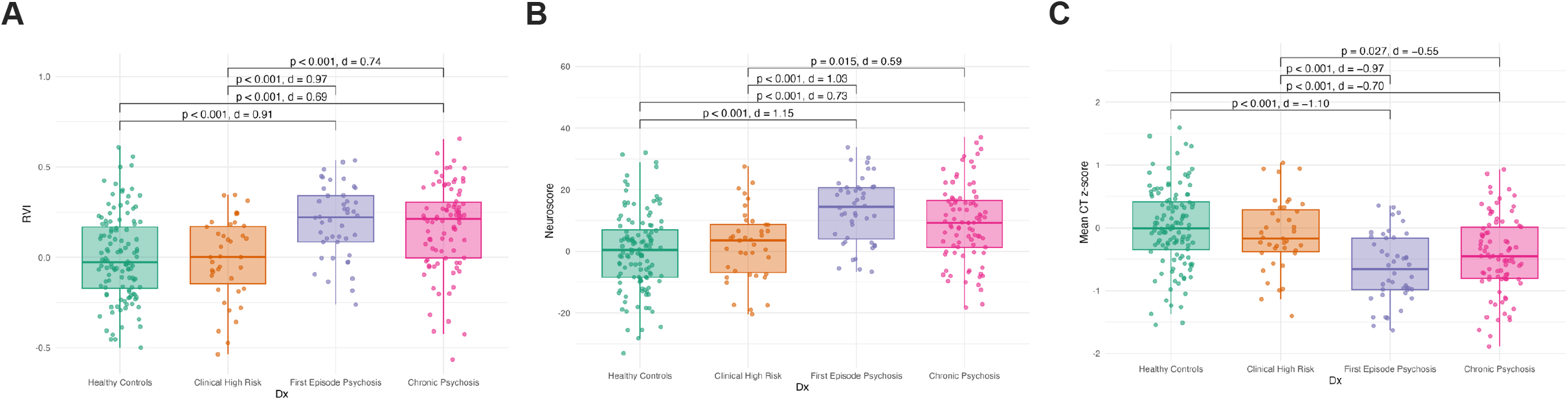
Group differences in structural neuroimaging indices across the early psychosis spectrum. Box plots showing the distribution of (**A**) the Regional Vulnerability Index (RVI), (**B**) the Neuroscore, and (**C**) the mean cortical thickness (CT) z-score in healthy controls (HC), clinical high-risk (CHR), first-episode psychosis (FEP), and chronic psychosis (CP) groups. All three indices differed significantly across the four groups. In post hoc comparisons, RVI and Neuroscore were significantly higher, and mean CT z-score significantly lower, in the FEP and CP groups relative to the CHR and HC groups; no significant differences were observed between FEP and CP or between CHR and HC. Boxes denote the interquartile range (IQR) with the median indicated by the horizontal line, whiskers extend to 1.5 × IQR. Brackets show Tukey-adjusted pairwise comparisons with corresponding p values and effect sizes (Cohen’s d).

Post hoc pairwise comparisons demonstrated that RVI and Neuroscore values were significantly higher in the FEP and CP groups than in the CHR and HC groups (RVI, all Tukey-adjusted *p*’s < .001, Cohen’s d = 0.69 to 0.97; Neuroscore, all Tukey-adjusted *p*’s < .05, Cohen’s d = 0.59 to 1.15), with no significant differences between FEP and CP or between CHR and HC (**Figure 1A, 1B**). Similarly, mean CT z-scores were significantly lower in the FEP and CP groups than in the CHR and HC groups (all Tukey-adjusted *p*’s < .05, Cohen’s d = −1.10 to −0.55), with no significant differences between FEP and CP or between CHR and HC (**Figure 1C**). Exact Tukey-adjusted *p* values and pairwise effect sizes (Cohen’s d) are presented in **Figure 1**.

### Associations among RVI, Neuroscore, and mean CT z-score

All three indices were significantly correlated with one another (**Figure 2, Table S4**). The strongest correlation was observed between Neuroscore and mean CT z-score (*r* = −0.997, *q* < .001), followed by Neuroscore and RVI (*r* = 0.624, *q* < .001), and RVI and mean CT z-score (*r* = −0.561, *q* < .001).

**Figure 2.**
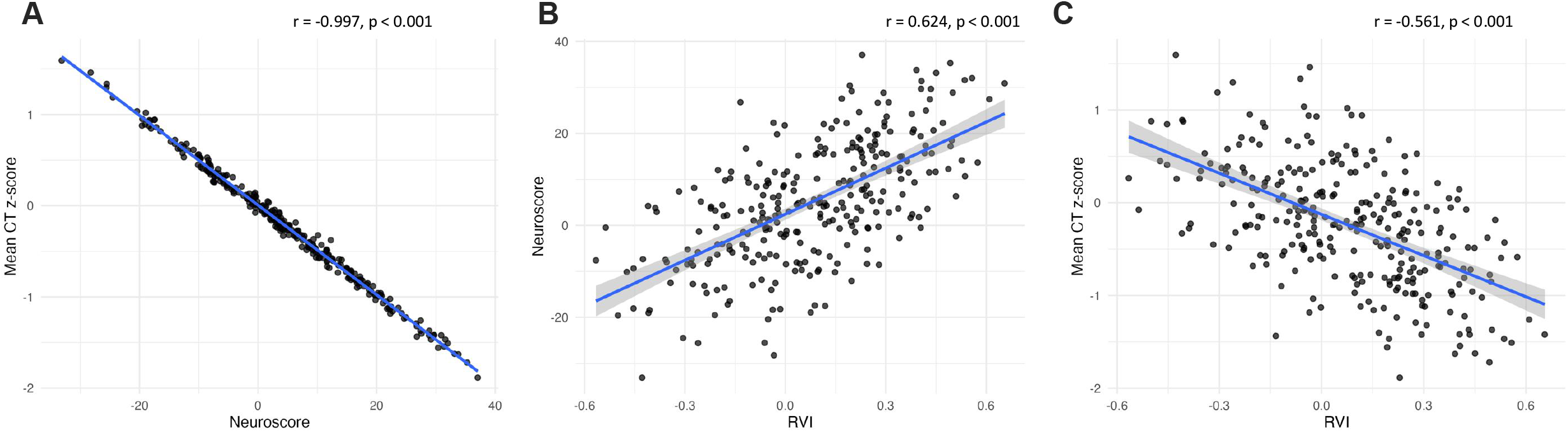
Pairwise associations among the three structural neuroimaging indices. Scatterplots with fitted linear regression lines (blue) and 95% confidence bands (shaded) showing the relationships between (**A**) Neuroscore and mean cortical thickness (CT) z-score, (**B**) the Regional Vulnerability Index (RVI) and Neuroscore, and (**C**) RVI and mean CT z-score. All three indices were significantly correlated with one another. Correlation coefficients (r) and FDR-adjusted p values for each pairwise association are reported on the figures.

### Associations between neuroimaging indices and clinical measures

None of the three indices (RVI, Neuroscore, or mean CT z-score) were significantly associated with with SIPS scores in CHR or PANSS scores in FEP or CP, either separately or combined, after FDR correction (**Table S6-S9**). In the combined psychosis sample (FEP and CP), longer disease duration was associated with lower Neuroscore (*r* = −0.270, *q* = .003) and higher mean CT z-score (*r* = 0.267, *q* = .003) (**Table S4**).

Several associations were significant but did not survive correction for multiple comparisons. In the CHR group, Neuroscore (*r* = −0.511, *p* = .025, *q* = .152) and mean CT z-score (*r* = 0.542, *p* = .017, *q*= .152) were significantly correlated with SIPS Negative scores. Additionally, RVI was positively correlated with PANSS Negative scores in the combined FEP and CP group (*r* = 0.203, *p* = .020, *q* = .246).

## DISCUSSION

In this study, we evaluated two individual-level neuroimaging biomarkers, CT-based RVI and Neuroscore, alongside mean CT deviation (z-score) across different stages of the psychosis spectrum, including CHR, FEP, and CP, and HC. RVI, Neuroscore, and mean CT z-score showed a consistent pattern across the psychosis spectrum: all three measures differed in individuals with established psychosis, including FEP and CP, compared with CHR and HC groups, but did not differ between FEP and CP or between CHR and HC. The high correlations among RVI, Neuroscore, and mean CT z-score suggest that these measures capture substantial shared variability in cortical thickness. None of the three measures were associated with SIPS symptom severity scores in CHR or PANSS symptom severity scores in FEP or CP. Together, these findings suggest that these CT-based individual-level biomarkers correspond more closely to established disease status than to risk status or current symptom burden in this cross-sectional sample, while their high intercorrelations underscore the need for more refined approaches that capture information beyond overall cortical thickness deviation.

These findings align with prior work indicating that both CT-based RVI and Neuroscore capture cortical alterations associated with established psychosis. Kochunov et al. reported significantly higher cortical RVI values in individuals with schizophrenia than in healthy controls.^9^ Consistent with our findings, cortical (CT-based) RVI did not differ across early, intermediate, and chronic stages of illness, whereas subcortical, white matter, and multimodal RVIs increased with illness progression. Similarly, Rodrigue et al. found higher CT-based Neuroscores in individuals with schizophrenia than in healthy controls.^10^ Our findings extend this work across distinct stages of psychotic illness, demonstrating comparable Neuroscore values in FEP and CP. Together, these results suggest that cortical RVI and Neuroscore capture disease-related cortical deviations that are evident in early course psychosis and show comparable values across illness stages in this cross-sectional sample.

Research examining RVI and Neuroscore in psychosis risk populations remains limited. Kochunov et al. reported that higher cortical (CT-based) RVI was associated with greater familial psychosis loading among 9-to 10-year-old children in the Adolescent Brain Cognitive Development (ABCD) Study cohort.^11^ However, cortical and other modality-based RVIs did not differ between children with and without a family history of psychosis. Also in the ABCD cohort, Karcher et al. found that compared with children in the low-distressing-PLE reference group, children with persistent, distressing PLEs showed elevated cortical RVIs, whereas no significant cortical RVI elevations were detected in the persistent-nondistressing, transient-distressing, or transient-nondistressing groups.^22^ Similarly, Cooper et al. reported higher CT-based Neuroscores among children with elevated and persistent distressing PLEs in the ABCD cohort.^23^ Our finding of no differences in CT-based RVI or Neuroscore between CHR and healthy control groups adds to this literature, suggesting that these measures may be more informative for identifying heterogeneity within risk populations than for distinguishing CHR status itself. The convergence of findings for CT-based RVI and Neuroscore is consistent with the strong correlation between these measures observed in our study and may reflect the substantial overlap in the cortical thickness information they capture.

Our finding of no difference in mean CT z-score between the CHR and HC groups is also consistent with prior work examining individual-level deviations in brain structure. In a large ENIGMA study of normative deviations in structural MRI measures among individuals at CHR, individual-level variation was largely nested within the healthy distribution and was not associated with positive symptom severity or subsequent conversion to psychosis.^7^

The high correlations among RVI, Neuroscore, and mean CT z-score may arise from their shared derivation from the same regional CT deviations and the widespread nature of CT alterations in schizophrenia. As a result, measures of similarity to the ENIGMA schizophrenia pattern also capture substantial information about overall CT deviation. The almost perfect correlation between mean CT z-score and Neuroscore is likely due to the limited variability and uniform direction of ENIGMA effect sizes (mean:-0.309, SD=0.098), together with the substantial intercorrelation of cortical thickness across regions. As a result, the weighted sum of CT values closely mirrors the unweighted mean. Future studies should determine whether this overlap extends to RVI and Neuroscore measures derived from other imaging modalities and multimodal approaches, which may capture complementary neurobiological information beyond cortical thickness.

We found no significant associations between psychosis symptom severity and CT-based measures. In the ENIGMA meta-analysis, positive and total PANSS scores were significantly correlated with regional thinner cortex, and higher negative PANSS scores were significantly correlated with widespread thinner cortex.^5^ Similarly, T1-weighted Neuroscore, which integrates cortical thickness, surface area, and subcortical volume measures, has been associated with higher positive and negative PANSS scores.^10^ RVI findings, on the other hand, have been mixed regarding symptom severity. Although prior work reports a significant correlation between RVI and the severity of negative symptoms, this association applies only to white matter RVI and multimodal RVI (a combination of cortical, subcortical, and white matter measures); other RVI modalities, such as cortical RVI, do not show this relationship.^9^ Although our moderate sample size may have limited our ability to detect smaller associations, together, our findings and previous studies provide converging evidence that biomarkers integrating information across multiple imaging measures or modalities may be more sensitive to symptom severity than CT-based measures alone.

Unexpectedly, longer disease duration was associated with higher mean CT z-scores and lower Neuroscore, indicating less CT deviation from healthy controls and lower similarity to ENIGMA schizophrenia-associated alterations, respectively. This finding should be interpreted cautiously given the strong relationship between disease duration and age. Age was regressed from cortical thickness measures in our analyses. However, age-related cortical changes have been shown to be non-linear, with a steeper decline up to the third decade of life and more gradual thereafter.^12^ Thus, linear adjustment for age could have contributed to the observed positive associations with disease duration. Longitudinal studies will be necessary to distinguish true illness-related cortical change from residual age and cohort effects.

Substantial within-group variability in RVI, Neuroscore, and mean CT z-score values was observed across the four study groups. Notably, some HC and CHR participants had higher RVI and Neuroscore values and lower mean CT z-scores compared to the mean levels observed in FEP and CP. This heterogeneity may limit the utility of these measures as stand-alone diagnostic or risk-classification tools at the individual level, even when significant group-level differences are present. At the same time, such variability may reflect meaningful biological heterogeneity within conventional diagnostic categories. Future work should therefore examine whether these measures are more informative when integrated with clinical, cognitive, genetic, or multimodal imaging features, or when used to identify biologically defined subgroups.

There are several limitations to consider when interpreting our findings. First, the cross-sectional nature of our analysis limits our ability to draw firm conclusions about stage-based differences in brain structure. Future longitudinal studies would help clarify the longitudinal trajectories of RVI and Neuroscore across different illness stages. Second, although our overall sample was reasonably large, subgroup sample sizes were modest and may have been underpowered for detecting some between-group differences. Third, detailed antipsychotic medication information and medication histories were not available for our sample. This limited our ability to address medication exposure as a potential confounder for the neuroimaging differences we observed. Fourth, both RVI and Neuroscore in this study were calculated using cortical thickness measures alone. Although prior studies have similarly used cortical-RVI, incorporating additional modalities such as subcortical volume, surface area, and white matter may yield findings of additional clinical importance. Future work incorporating these modalities may detect symptom-domain associations that our cortical-only approach was unable to capture.

In our cohort spanning healthy controls, clinical high risk, first-episode psychosis, and chronic schizophrenia, RVI, Neuroscore, and mean CT z-score were markers of established disease status rather than risk status or illness stage. Their high intercorrelations suggest that these measures capture substantial shared variability in cortical thickness, while the marked within-group variability across diagnostic groups highlights a barrier to their clinical translation. Nevertheless, these findings support the promise of RVI and Neuroscore as individualized neuroimaging approaches and underscore the need for further refinement through data-driven methods that do not depend on meta-analytic group-level findings, as well as multimodal methods that capture complementary neurobiological information.

## Data Availability

All data produced in the present study are available upon reasonable request to the authors

