## Supplemental Tables for "Individual-level Structural Neuroimaging Biomarkers across the Psychosis Spectrum"

**SUPPLEMENTARY MATERIALS**

**Supplementary Table S1. Scanner distribution by diagnostic groups**

| **Dx** | **Prisma** | **TrioTim** |
| --- | --- | --- |
| HC | 68 | 52 |
| CHR | 39 | 0 |
| FEP | 37 | 7 |
| CP | 36 | 50 |

**Supplementary Table S2. MRI Scanner Parameters and Participant Demographics**

| **Scan Parameters** | **Siemens MAGNETOM Prisma** | **Siemens Tim Trio** |
| --- | --- | --- |
| Field Strength | 3T | 3T |
| Head Coil | 64-channel | 32-channel |
| Sequence | TI-weighted MPRAGE | T1-weighted MPRAGE |
| Voxel Size (mm') | 1.0 × 1.0 ×1.0 | 1.0x1.0x1.0 |
| TR (ms) | 2530 | 1900 |
| TE (ms) | 1.69 | 2.26 |
| FOV (mm) | 256 | 250 |
| Number of Slices | 176 | 17 |

**Supplementary Table S3. Scanner comparisons of RVI, Neuroscore, and mean CT z-score within diagnostic groups**

| **Dx** | **Measure** | ***t*** | ***df*** | ***p*** | **FDR-adjusted *p*** |
| --- | --- | --- | --- | --- | --- |
| FEP | RVI | -0.165 | 7.864 | 0.873 | 0.994 |
| FEP | Neuroscore | 0.008 | 7.464 | 0.994 | 0.994 |
| FEP | Mean CT z-score | -0.083 | 7.359 | 0.936 | 0.994 |
| CP | RVI | -0.276 | 72.946 | 0.783 | 0.783 |
| CP | Neuroscore | -1.098 | 76.197 | 0.276 | 0.413 |
| CP | Mean CT z-score | 1.142 | 76.755 | 0.257 | 0.413 |
| HC | RVI | 1.192 | 106.135 | 0.236 | 0.396 |
| HC | Neuroscore | 0.903 | 105.704 | 0.368 | 0.396 |
| HC | Mean CT z-score | -0.852 | 106.800 | 0.396 | 0.396 |

**Supplementary Table S4. Core neuroimaging measure correlations (All participants)**

| **Neuroimaging Measure** | **Clinical Measure** | **n** | **Correlation Coefficient (*r*)** | ***p*** | **FDR-adjusted *p*** |
| --- | --- | --- | --- | --- | --- |
| RVI | Neuroscore | 289 | 0.624 | <2.2e-16 | <2.2e-16 |
| RVI | Mean CT z-score | 289 | -0.561 | <2.2e-16 | <2.2e-16 |
| Neuroscore | Mean CT z-score | 289 | -0.997 | <2.2e-16 | <2.2e-16 |

**Supplementary Table S5. Correlations with disease duration (FS+SZ)**

| **Neuroimaging Measure** | **n** | **ρ** | ***p*** | **FDR-adjusted *p*** |
| --- | --- | --- | --- | --- |
| RVI | 130 | -0.120 | .174 | .174 |
| Neuroscore | 130 | -0.270 | .002 | .003 |
| Mean CT z-score | 130 | 0.267 | .002 | .003 |

**Supplementary Table S6. Correlations with SIPS measures (CHR)**

| **Neuroimaging Measure** | **Clinical Measure** | **n** | **Correlation Coefficient (*r*)** | ***p*** | **FDR-adjusted *p*** |
| --- | --- | --- | --- | --- | --- |
| RVI | SIPS Negative | 19 | 0.028 | .908 | .908 |
| Neuroscore | SIPS Negative | 19 | -0.511 | .025 | .152 |
| Mean CT z-score | SIPS Negative | 19 | 0.542 | .017 | .152 |
| RVI | SIPS General | 20 | 0.073 | .759 | .908 |
| Neuroscore | SIPS General | 20 | -0.032 | .894 | .908 |
| Mean CT z-score | SIPS General | 20 | 0.036 | .879 | .908 |
| RVI | SIPS Total | 18 | 0.029 | .908 | .908 |
| Neuroscore | SIPS Total | 18 | -0.399 | .101 | .303 |
| Mean CT z-score | SIPS Total | 18 | 0.422 | .081 | .303 |
| RVI | SIPS Positive | 36 | -0.140 | .414 | .908 |
| Neuroscore | SIPS Positive | 36 | -0.080 | .641 | .908 |
| Mean CT z-score | SIPS Positive | 36 | 0.076 | .660 | .908 |

**Supplementary Table S7. Correlations with PANSS measures (FEP+CP)**

| **Neuroimaging Measure** | **Clinical Measure** | **n** | **Correlation Coefficient (ρ)** | ***p*** | **FDR-adjusted *p*** |
| --- | --- | --- | --- | --- | --- |
| RVI | PANSS Total | 130 | 0.125 | .155 | 0.746 |
| Neuroscore | PANSS Total | 130 | 0.073 | .410 | 0.819 |
| Mean CT z-score | PANSS Total | 130 | -0.055 | .534 | 0.838 |
| RVI | PANSS General | 130 | 0.014 | .872 | 0.998 |
| Neuroscore | PANSS General | 130 | 0.009 | .915 | 0.998 |
| Mean CT z-score | PANSS General | 130 | -0.0002 | .998 | 0.998 |
| RVI | PANSS Negative | 130 | 0.203 | .021 | 0.246 |
| Neuroscore | PANSS Negative | 130 | 0.052 | .559 | 0.838 |
| Mean CT z-score | PANSS Negative | 130 | -0.037 | .676 | 0.902 |
| RVI | PANSS Positive | 130 | 0.114 | .198 | 0.746 |
| Neuroscore | PANSS Positive | 130 | 0.102 | .249 | 0.746 |
| Mean CT z-score | PANSS Positive | 130 | -0.083 | .349 | 0.819 |

**Supplementary Table S8. Correlations with PANSS measures (FEP only)**

| **Neuroimaging Measure** | **Clinical Measure** | **n** | **Correlation Coefficient (ρ)** | ***p*** | **FDR-adjusted *p*** |
| --- | --- | --- | --- | --- | --- |
| RVI | PANSS Total | 44 | 0.146 | .344 | .516 |
| Neuroscore | PANSS Total | 44 | 0.233 | .128 | .452 |
| Mean CT z-score | PANSS Total | 44 | -0.203 | .185 | .452 |
| RVI | PANSS General | 44 | 0.040 | .794 | .794 |
| Neuroscore | PANSS General | 44 | 0.176 | .253 | .452 |
| Mean CT z-score | PANSS General | 44 | -0.172 | .263 | .452 |
| RVI | PANSS Negative | 44 | 0.278 | .068 | .452 |
| Neuroscore | PANSS Negative | 44 | 0.117 | .450 | .540 |
| Mean CT z-score | PANSS Negative | 44 | -0.087 | .575 | .627 |
| RVI | PANSS Positive | 44 | 0.122 | .430 | .540 |
| Neuroscore | PANSS Positive | 44 | 0.247 | .106 | .452 |
| Mean CT z-score | PANSS Positive | 44 | -0.200 | .193 | .452 |

**Supplementary Table S9. Correlations with PANSS measures (CP only)**

| **Neuroimaging Measure** | **Clinical Measure** | **n** | **Correlation Coefficient (ρ)** | ***p*** | **FDR-adjusted *p*** |
| --- | --- | --- | --- | --- | --- |
| RVI | PANSS Total | 86 | 0.131 | .228 | .912 |
| Neuroscore | PANSS Total | 86 | 0.041 | .710 | .988 |
| Mean CT z-score | PANSS Total | 86 | -0.012 | .911 | .988 |
| RVI | PANSS General | 86 | 0.008 | .945 | .988 |
| Neuroscore | PANSS General | 86 | -0.043 | .697 | .988 |
| Mean CT z-score | PANSS General | 86 | 0.064 | .555 | .988 |
| RVI | PANSS Negative | 86 | 0.150 | .169 | .912 |
| Neuroscore | PANSS Negative | 86 | 0.002 | .988 | .988 |
| Mean CT z-score | PANSS Negative | 86 | 0.016 | .887 | .988 |
| RVI | PANSS Positive | 86 | 0.138 | .204 | .912 |
| Neuroscore | PANSS Positive | 86 | 0.112 | .306 | .919 |
| Mean CT z-score | PANSS Positive | 86 | -0.088 | .421 | .988 |
